# A survey of Large Language Model use in a hospital, research, and teaching campus

**DOI:** 10.1101/2024.09.11.24313512

**Authors:** Loretta Gasparini, Nitya Phillipson, Daniel Capurro, Revital Rosenberg, Jim Buttery, Jayne Howley, Sarath Ranganathan, Catherine Quinlan, Niloufer Selvadurai, Michael Wildenauer, Michael South, Gerardo Luis Dimaguila

## Abstract

**Background:** The use of Large Language Models (LLMs) has exploded since November 2022 but there is sparse evidence regarding LLM use in health, medical and research contexts.

**Objective:** To summarise the current uses of and attitudes towards LLMs across the clinical, research and teaching contexts in our campus.

**Design:** We administered a survey about LLM uses and attitudes. We conducted summary quantitative analysis and inductive qualitative analysis of free text responses.

**Setting:** In August-September 2023, we circulated the survey amongst all staff and students across our campus (approximately n=7500), a fully integrated paediatric academic hospital and research institute.

**Participants:** We received 281 anonymous survey responses.

**Main outcome measures:** We asked about participants’ knowledge of LLMs, their current use of LLMs in professional or learning contexts, and perspectives on possible future uses, opportunities, and risks of LLM use.

**Results:** Over 90% of respondents have heard of LLM tools and about two-thirds have used them in their work on our campus. Respondents reported using LLMs for a range of uses, including for generating or editing text and exploring ideas. Many, but not necessarily all, respondents seem aware of the limitations and potential risks of LLMs, including privacy and security risks. Various respondents expressed enthusiasm about opportunities of LLM use, including increased efficiency.

**Conclusions:** Our findings show LLM tools are already widely used on our campus. Guidelines and governance are needed to keep up with practice. We have developed recommendations for the use of LLMs on our campus using insights from this survey.

**What is known:** **The known:** The use of Large Language Models (LLMs) has increased rapidly since the introduction of ChatGPT in November 2022.

**The new:** Most survey respondents are aware of, if not using, LLMs in their work across our hospital, research, and university campus. Diverse uses were reported, including generating or editing text and exploring ideas. There were varying attitudes towards LLMs. Perceived risks included privacy and security risks. A key perceived opportunity was increased efficiency.

**The implications:** LLM tools are already widely used on our campus, highlighting the need for guidelines and governance to keep up with practice.

## 1. Introduction

Large Language Models (LLMs) are computational models trained on huge amounts of text to recognize and mimic nuanced patterns in human language, allowing them to receive multimodal prompts and generate responses approximating human performance on many tasks (1,2). The performance, public availability and awareness of LLMs increased dramatically after the introduction of ChatGPT in November 2022 (1), leading to extensive use in professional and learning settings. This has led to unprecedented challenges in ensuring LLMs are used in responsible and ethical ways (e.g., (3,4)), accentuated by their typically opaque nature and frequently undocumented use in workplaces (5). While the use of these tools presents opportunities for efficiencies and innovations in administrative, clinical and research settings, concerns about clinical, research and education applications of LLMs include confidentiality and privacy concerns, inaccuracies, hallucinations (generation of content that is not real), medical liability, biases embedded within models and a lack of accountability or transparency in how LLMs make decisions (6–8).

The Melbourne Childen’s Campus is a fully integrated paediatric academic hospital and research institute. While anecdotally it appeared LLMs were used frequently throughout the Campus in clinical, research and teaching contexts, we did not previously have evidence of how prevalent this was and in exactly which contexts and for what purposes LLMs were used. We also did not have evidence of the attitudes towards LLMs in our contexts. Furthermore, we wished to understand whether the domains of clinical care, research and teaching presented any differences or similarities in relation to LLM use, risks and opportunities. Such evidence would help support better governance of LLM and broader Generative Artificial Intelligence (GenAI) use and integration across our Campus. Whilst expert viewpoints and reviews on LLM use and opportunities have been reported for medical education (9–12), drug discovery, (13,14) and higher degree research (15), there is sparse evidence for them in the context of an integrated specialist paediatric academic hospital and research campus.

### 1.1. Objective

We aimed to summarise the current uses and attitudes towards LLMs in our Campus across the clinical, research and teaching contexts.

## 2. Methods

### 2.1. Context

Our Campus spans a paediatric hospital, children’s medical research institute, and paediatric university department in a metropolitan, Australian city. The paediatric hospital is a quaternary hospital with over 6000 staff and 350 beds. The medical research institute employs over 1800 researchers working across 150 diseases and conditions affecting children. The university paediatric department comprises over 70 academic staff, 15 professional staff, 150 graduate research students, and over 400 Honorary staff members. We estimate there are approximately 7500 staff and students across the Campus, although, many have joint affiliations across the three partners.

### 2.2. Qualitative approach

Based on our objective of generating insights from participants across the Campus, we followed a systematic grounded theory approach. We circulated a survey amongst all staff and students across our Campus to gather information about current uses, expected future uses, opportunities, and risks of LLM use in clinical, research, and teaching and learning contexts. The survey is available in Appendix 1. As most questions on LLM use and attitudes were open-ended, we followed the Standards for Reporting Qualitative Research (SRQR) (16).

The researchers involved in the survey design, data collection, and data analysis were comprised of a Working Group assembled to assess the current state of LLM use in academic child health and identify opportunities and challenges for the future. The group’s positions and expertise are broad, spanning early and senior researchers, clinicians, informaticians and data scientists, and leadership, as well as child clinical care consumer representatives.

### 2.3. Participants

All staff and students across our Campus were eligible to participate. This spans clinicians, health and medical researchers, postgraduate researchers, data scientists and administrative and support staff.

This quality improvement project received quality assurance approval from the Royal Children’s Hospital Research Ethics & Governance Office (100638).

### 2.4. Survey design

We developed a survey based on an iterative discussion between members of the Working Group. MS drafted survey items based on the group discussion. It was then piloted with the Working Group and refined until the group was satisfied. During this process we referred to a survey of GenAI use in commercial entities (17).

### 2.5. Data collection methods

We created a web-based survey using LimeSurvey (https://www.limesurvey.org/). We distributed the survey via various mailing lists across the Campus, estimated to have reached about 1000 clinical staff, 1800 researchers and 100 students. The survey was open for a period of 4 weeks (28^th^ August to 22^nd^ September 2023).

### 2.6. Data processing and analysis

Data were deidentified and aggregated and distinctive written text paraphrased to avoid identification. We performed statistical analysis of descriptive and quantitative data using Microsoft Excel. We did not perform any inferential statistics.

We analysed free text responses using inductive analysis in NVivo 14 (18). The data were coded evenly by GLD and LG. First, GLD coded a segment of the data and established initial codes and definitions. The remaining data were single coded by GLD or LG, who added new codes and definitions as needed. The definitions of codes were refined by discussion. After coding, GLD conducted an inductive thematic analysis of the codes. The resulting themes were reviewed and approved by LG and NP.

For the purposes of establishing a timely overview of the current uses and perceptions of LLM use in our campus, we deemed it sufficient to single code the data.

## 3. Results

### 3.1. Summary statistics

We received 281 survey responses. Most respondents had an affiliation with the hospital (n=174), many with the research institute (n=131) and fewer with the university department (n=85) (many respondents have more than one affiliation, so these numbers exceed total respondents). Over half reported having a clinical role (n=158), half in research (n=140), about one quarter in teaching (n=75) and a tenth in data services (n=27) (again, respondents can work in multiple capacities).

Respondents were asked to report what best described their level of experience. Most identified as ‘senior’ (senior, fully qualified, post-doctoral, senior data analyst or developer; n=201) and fewer as ‘junior’ (junior, trainee, student, junior data analyst or developer; n=56).

### 3.2. Quantitative results

Over 90% of staff were familiar with LLM tools, most commonly ChatGPT. Regarding use of LLM tools, 64% reported current or previous use of any LLM tool in their work. Figure 1 shows respondents’ reported familiarity with and use of specific LLM tools. The vast majority was ChatGPT (GPT3, 63%; GPT4, 19%). There was also some use of Grammarly (24%) and Microsoft Bing Chat (7%) and very small numbers reported using any other tools.

**Figure 1.**
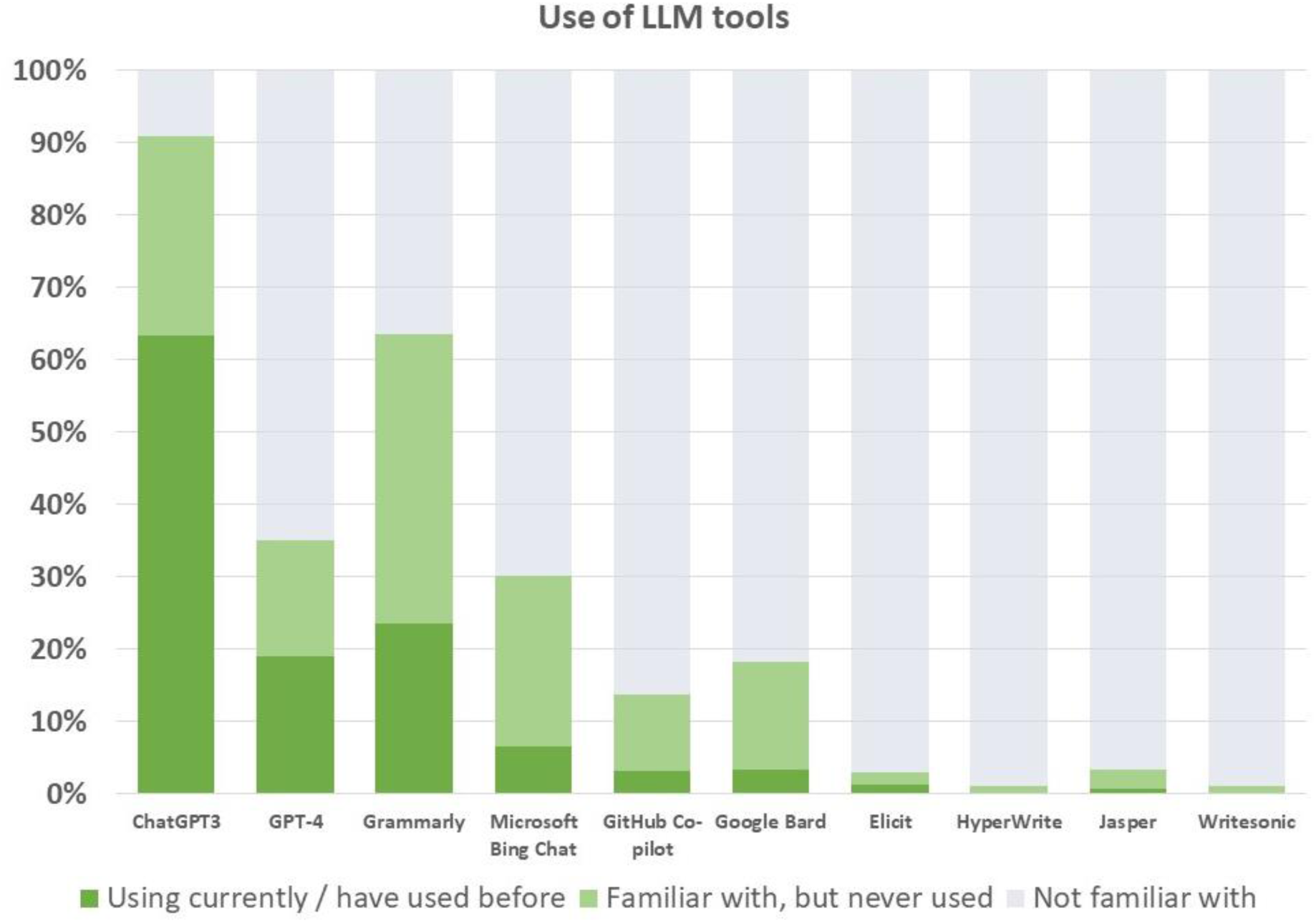
Reported familiarity with and use of LLMs. Dark represents use of the LLM tool, light green familiarity with but no use of the tool, and grey no use of the tool

In addition, some people reported having heard of but never using ChatGPT (GPT3, 28%; GPT4, 16%) or Grammarly (40%). Some had heard of but never used Microsoft Bing Chat (23%), Google Bard (15%) or GitHub Co-pilot (10%). Very small numbers reported familiarity with other tools.

### 3.3. Qualitative synthesis

The responses suggest a wide array of current LLM uses in clinical and research settings. Respondents seemed mostly very aware of potential benefits of the use of LLMs but also of a range of possible risks.

#### 3.3.1. Current and future uses

A substantial number of respondents reported using LLMs for generating content (e.g., generating or editing text). See Table 1 for example quotes for the themes we describe here. Other uses included knowledge support (e.g., exploring ideas, identifying areas of improvement or help writing programming code), or data processing (e.g., data analysis, extraction, management, or interpretation) and using LLM as an information source or to find information sources. Relatively few reported using LLMs for Natural Language Processing or administrative tasks. Various respondents also reported no current use of LLMs, as reflected by the quantitative results.

**Table 1.** Example quotes from key themes of current and future uses of LLMs.

| Theme: Code | Quote (career level, affiliations) |
| --- | --- |
| **Current uses** |  |
| Content generation: Text generation, research support | *"I have used them to summarise research papers"*<br>(junior; research) |
|  | *"generate titles for articles/abstracts/presentations"*<br>(senior; research) |
| Knowledge Support: Clinical Decision Support | *"Symptom/sign input to generate a list of differential diagnoses"* (junior; clinical, research, teaching & learning) |
| Data Processing: | *"As English is my second language, it helps me enhance the clarity and professionalism of my notes and letters"*<br>(senior; clinical) |
| Information Source: Seek info sources | *"helped in getting information sources rather than get the information itself"* (junior; clinical, teaching & learning) |
| **Future uses** |  |
| Content Generation: Text generation, comms | *"Translation of clinic letters/fact sheets into other languages"* (junior; clinical) |
| Content Generation: Text generation, research support | *"Haven't used anything yet but can envisage using AI for first drafts for literature reviews, consents, reports"*<br>(senior; research) |
| Content Generation: Text generation, teaching activities | *"Generation of learning materials on specific topics"*<br>(senior; clinical, teaching) |
| Knowledge Support: Clinical Decision Support | *"Could also be used in triaging patients - entering in symptoms/signs etc to expedite accurate triage prior to clinical assessment"* (senior; clinical) |
| Data Processing: Data Analysis from Queries | *"analysis of large data sets to look for answers to science / health / research questions to support policy changes and implementation, new / updated clinical practices, new treatment options, new diagnosis,* |
|  | *understanding disease conditions, etc”* (senior; research) |
| Knowledge Support: Computer Programming | *“Help with data analysis and syntax for statistical programs”* (junior; research, teaching & learning) |
| Increased Efficiency: Grading and reviewing | *“help with grading”* (senior; teaching) |
| Knowledge Support: Human skills | *“students will use it, so we need to make sure we train them how to. Train how to be critical users”* (senior; teaching) |

Respondents frequently identified potential future uses related to generating text, which matches currents uses of LLM. Respondents also frequently foresaw that LLMs could be used in future for clinical decision support, research activities like data analysis or computer programming and teaching activities like generating learning materials, grading and reviewing and developing human skills.

#### 3.3.2. Opportunities and risks

The primary opportunity identified by respondents was efficiency, for instance, saving time or requiring few resources. See Table 2 for example quotes for the themes we describe here. Some respondents saw opportunities to expand human skills using LLMs. Some reported that using LLMs could result in higher quality and more accurate outputs, although there were more reports of the risk of *lower* quality and accuracy.

**Table 2.**
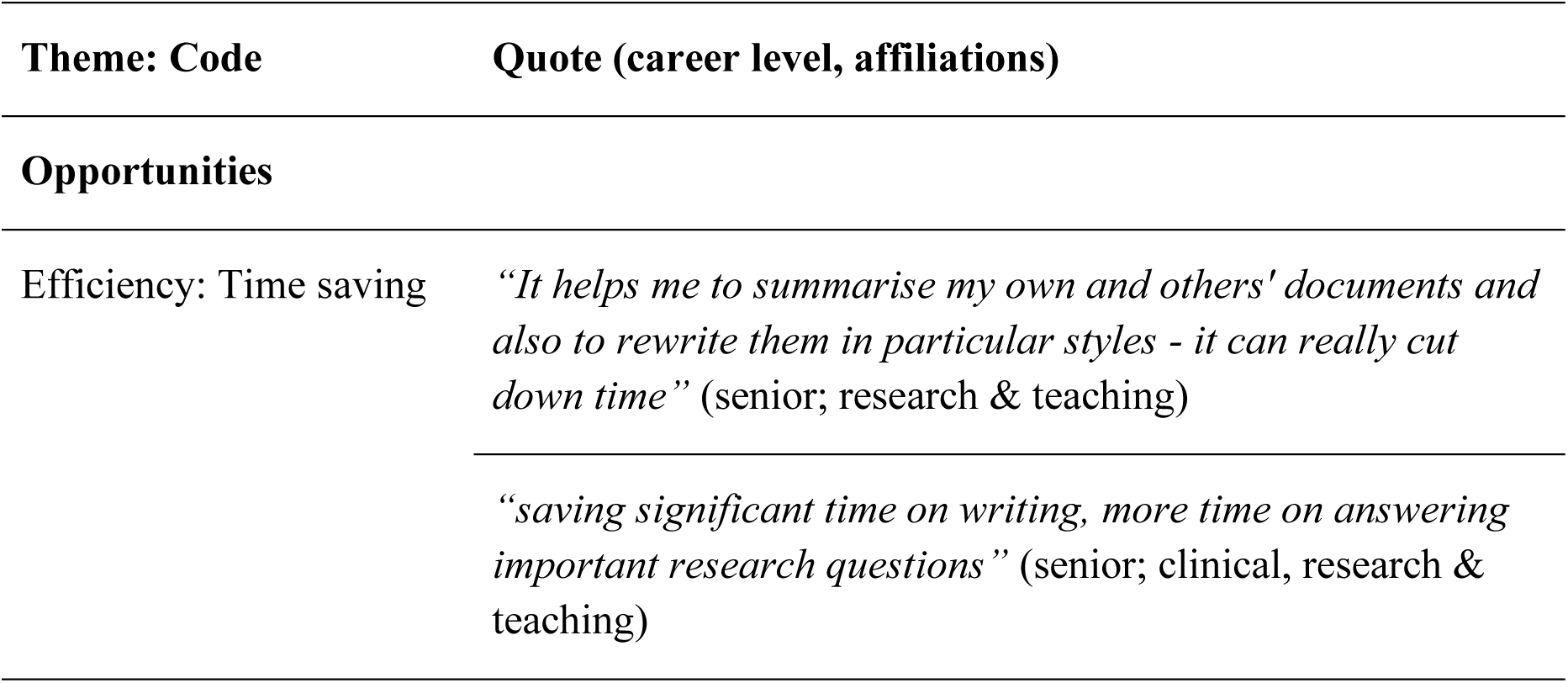
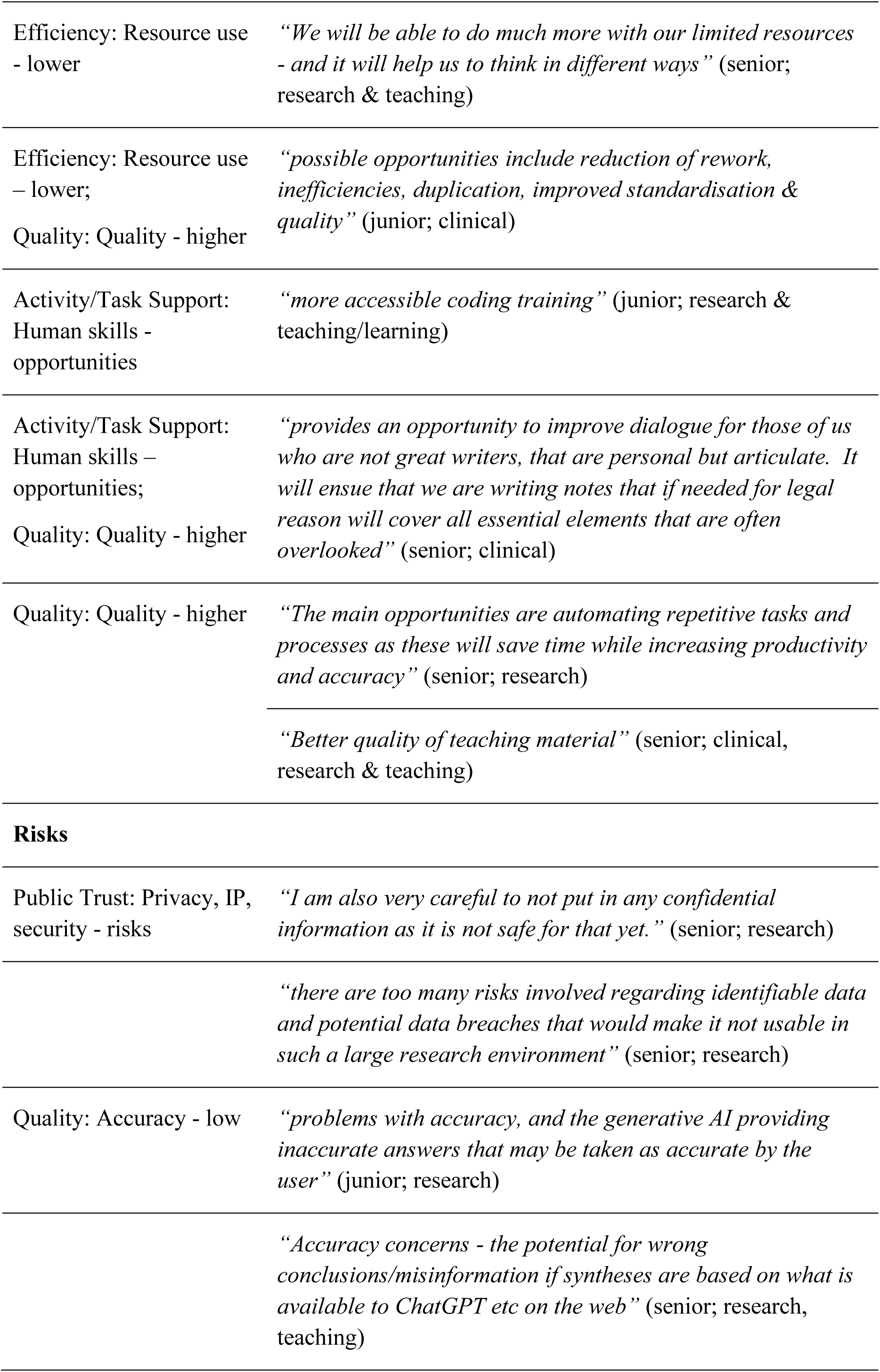
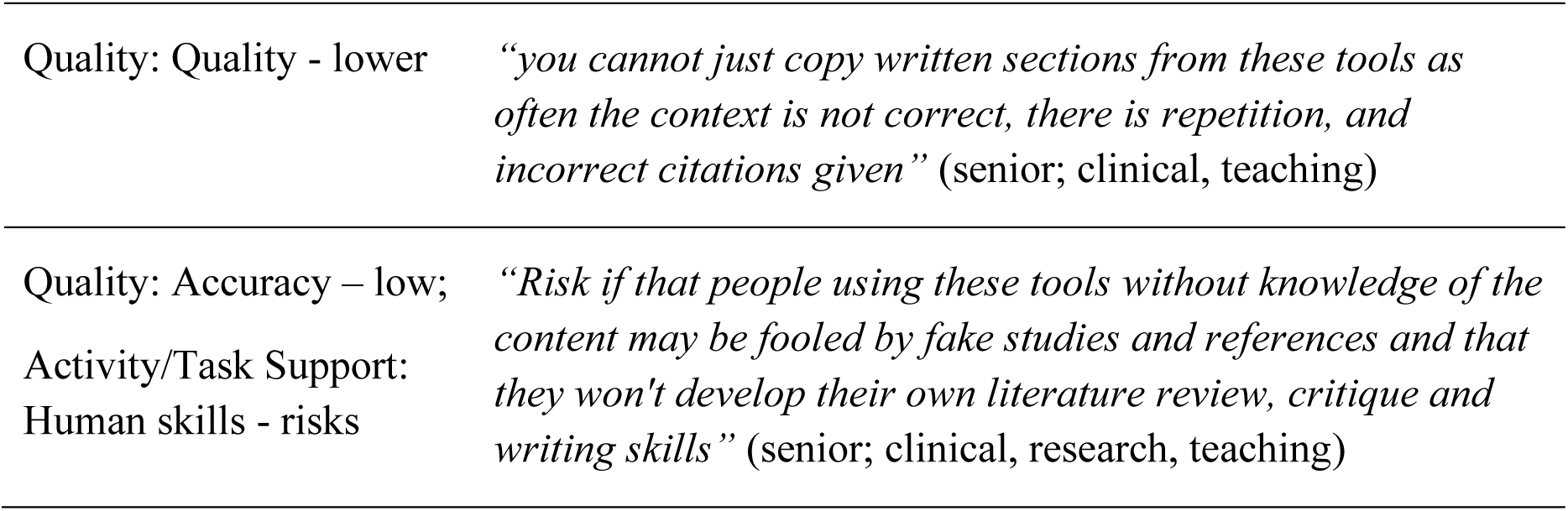
Example quotes from key themes of opportunities and risks of LLMs.

| Theme: Code | Quote (career level, affiliations) |
| --- | --- |
| **Opportunities** |  |
| Efficiency: Time saving | *“It helps me to summarise my own and others' documents and also to rewrite them in particular styles - it can really cut down time”* (senior; research & teaching) |
|  | *“saving significant time on writing, more time on answering important research questions”* (senior; clinical, research & teaching) |
| Efficiency: Resource use<br>- lower | *“We will be able to do much more with our limited resources*<br>*- and it will help us to think in different ways”* (senior;<br>research & teaching) |
| Efficiency: Resource use<br>– lower;<br>Quality: Quality - higher | *“possible opportunities include reduction of rework,*<br>*inefficiencies, duplication, improved standardisation &*<br>*quality”* (junior; clinical) |
| Activity/Task Support:<br>Human skills -<br>opportunities | *“more accessible coding training”* (junior; research &<br>teaching/learning) |
| Activity/Task Support:<br>Human skills –<br>opportunities;<br>Quality: Quality - higher | *“provides an opportunity to improve dialogue for those of us*<br>*who are not great writers, that are personal but articulate. It*<br>*will ensue that we are writing notes that if needed for legal*<br>*reason will cover all essential elements that are often*<br>*overlooked”* (senior; clinical) |
| Quality: Quality - higher | *“The main opportunities are automating repetitive tasks and*<br>*processes as these will save time while increasing productivity*<br>*and accuracy”* (senior; research) |
|  | *“Better quality of teaching material”* (senior; clinical,<br>research & teaching) |
| **Risks** |  |
| Public Trust: Privacy, IP,<br>security - risks | *“I am also very careful to not put in any confidential*<br>*information as it is not safe for that yet.”* (senior; research) |
|  | *“there are too many risks involved regarding identifiable data*<br>*and potential data breaches that would make it not usable in*<br>*such a large research environment”* (senior; research) |
| Quality: Accuracy - low | *“problems with accuracy, and the generative AI providing*<br>*inaccurate answers that may be taken as accurate by the*<br>*user”* (junior; research) |
|  | *“Accuracy concerns - the potential for wrong*<br>*conclusions/misinformation if syntheses are based on what is*<br>*available to ChatGPT etc on the web”* (senior; research,<br>teaching) |
| Quality: Quality - lower | *“you cannot just copy written sections from these tools as often the context is not correct, there is repetition, and incorrect citations given”* (senior; clinical, teaching) |
| Quality: Accuracy – low;<br>Activity/Task Support:<br>Human skills - risks | *“Risk if that people using these tools without knowledge of the content may be fooled by fake studies and references and that they won't develop their own literature review, critique and writing skills”* (senior; clinical, research, teaching) |

The main risks identified related to privacy, intellectual property (IP), security, integrity and public trust, particularly by clinical or research respondents. Low accuracy was also a common risk reported by respondents across all three affiliations. Many teaching and learning respondents (i.e. educators or students) indicated their concern that LLM outputs are likely to be of lower quality.

After these risks, other risk themes were identified infrequently but included the risk of misinterpreting or uncritically using LLM output, integrity risks, low transparency and oversight for appropriately using LLMs and loss of human skills when relying on LLMs.

## 4. Discussion

### 4.1. Main findings

We found most respondents have heard of LLM tools and about two-thirds have used them in their work. Respondents reported using LLMs for various uses, including generating or editing text, knowledge support, data processing and as a source of information. Many respondents identified a major opportunity of LLM use as increased efficiency. Many respondents seemed aware and realistic about the limitations and potential risks of LLMs. Privacy, IP and security concerns were the most frequently cited risks of LLM use, as well as lower accuracy and quality of outputs generated by LLMs. In contrast, some of the respondents’ comments suggest they may not be aware of these risks, particularly the entering of private information into LLMs or relying on LLMs as an information source when they are not verified to be reliable for this purpose (e.g., see example quotes in Table 1). This suggests the need for clearer and more accessible usage guidelines and training. A few themes arose both as an opportunity and risk (e.g., higher or lower accuracy and quality, or the increase or decrease of human skills), highlighting that opinions differ on LLM capabilities and potential.

Many of the respondents’ attitudes echo issues reported in the literature around LLM use in medical education and higher degree research. Some key opportunities identified in a systematic review of LLM use in medical education were for creating courses and assessment, using LLMs as a writing tool, and allowing greater access to published knowledge and research (12). These correspond to the many reports of current use of LLMs in the themes of content generation and knowledge support. The key challenges the authors of that review identified were ethical, legal and privacy, academic integrity, incorrect responses and overreliance. These correspond to the risks reported by our respondents around privacy, IP, security, integrity and public trust, accuracy and human skills. Other viewpoint and editorial papers on LLM use in medical research have expressed similar attitudes on opportunities and risks (e.g., (9,10). A critical dialogue between two academics in higher education also highlights the tension between using tools to improve efficiency whilst retaining academic integrity (15).

### 4.2. Implications

Our findings show that LLM tools are already widely used on our campus and for a range of uses. We were concerned by some of the uses identified by respondents that implied potential confidentiality and privacy breaches, which would violate the Victorian Department of Health’s advice on use of unregulated Artificial Intelligence (AI) in health services (19). Certain responses also implied some respondents may use LLMs as an information source, when they are not reliable for this purpose. This only strengthens the pressing need to develop agile policies and relevant training that evolves with emerging use cases and resultant risks. Thus, members of our Working Group drafted recommendations for the use of LLMs and GenAI more broadly on our campus (see Appendix 2).

We consider banning the use of LLM tools in certain domains is unlikely to work in practice. These tools are already getting significant use and users are experiencing obvious benefits, including from major electronic medical record providers in Australia (20). There is no practical way to limit access and the very same tools are publicly available to patients and their families.

It is also important to recognise the enthusiasm and interest staff have around the benefit of LLMs, and encourage them to explore the technology, as long as this is done with transparency for the organisations and with appropriate governance informed by local and external policies.

To ensure LLM use leads to higher accuracy and quality of outputs and expansion of human skills, users could be provided training and resources on effective LLM use, for example on prompt engineering strategies and critical appraisal of LLM output. Such resources should also provide guidance for ethical and responsible use of LLMs.

Beyond contributing to the formulation of internal guidelines and policies, it is hoped that the findings of our survey will support the federal and state governments’ work in this area, such as the 2024 Safe and Responsible AI in Australia consultation (21). Here, the government’s recommended ‘risk-based approach’ to regulation requires lawmakers to identify low- and high-risk AI applications in various sectors. Thus, the spectrum of activities identified in our survey can provide insights on the nature and extent of LLM use in the health sector. Given the rapid evolution of AI tools, we suspect developing generalisable principles for responsible AI use may be more sustainable than overly specific rules.

### 4.3. Transferability

We expect the exact uses of LLM tools will differ across different institutions, but the results of the current study may reflect general patterns in academic, clinical and tertiary education contexts. Other institutions may repeat our methods to better understand the use of LLMs in their context and should develop guidelines relevant to their context.

### 4.4. Limitations

Our survey was completed voluntarily across our Campus, so the quantitative results represent LLM use of respondents, not across the entire Campus. Our qualitative results provide a summary of uses of and attitudes towards LLMs on our campus but are not representative of all our staff and students. Our survey responses also reflect a snapshot in time (August-September 2023) of LLM uses and attitudes, which are expected to change as the landscape of LLMs is rapidly developing. However, the results provide us with a baseline understanding of LLMs use and attitudes in our context and will allow us to track changes over time.

## 5. Conclusion

We surveyed the current uses and attitudes towards large language models in our campus across clinical, research and teaching contexts. Most respondents have used LLMs in their work for a range of uses. This highlights the need for governance to keep up with practice. We have used the insights gained through this survey to develop recommendations for the use of GenAI and LLMs on our campus.

## Authors’ Note

### Citation

Gasparini, L., Phillipson, N., Capurro, D., Rosenberg, R., Buttery, J., Howley, J., Ranganathan, S., Quinlan, C., Selvadurai, N., Wildenauer, M., South, M., Dimaguila, GL. (2024). A survey of Large Language Model use in a hospital, research, and teaching campus.

### Preprint

This preprint has not yet been peer reviewed.

### Author contributions

MS drafted the data collection survey and conducted the quantitative analysis, GLD and LG coded the qualitative data, GLD conducted the qualitative analysis, LG drafted the original manuscript, all authors approved the data collection survey and conceptualized, reviewed, edited, and approved the final manuscript.

### Ethics statement

This quality improvement project received quality assurance approval from the Royal Children’s Hospital Research Ethics & Governance Office (100638).

### Funding

LG was supported by an Australian Government Research Training Program (RTP) Scholarship and MCRI PhD Top Up Scholarship. Research at the Murdoch Children’s Research Institute (MCRI) was supported by the Victorian Government’s Operational Infrastructure Support Program. The funding organizations are independent of all researchers and were not involved in any of the study design, the collection, analysis, and interpretation of data, the writing of the report or the decision to submit the manuscript for publication.

### Data access

The authors had full access to all the data (including statistical reports and tables) in the study.

### Survey availability

The survey used for data collection is available in Appendix 1.

### Data availability statement

The data that support the findings of this study are available upon reasonable request from the corresponding author, LG. The data are not publicly available due to privacy of the participants.

## Data Availability

The survey used for data collection is available in Appendix 1. The data that support the findings of this study are available upon reasonable request from the corresponding author, LG. The data are not publicly available due to privacy of the participants.

## Acknowledgements

We thank all the respondents to our survey for their time and contribution. For insightful contributions, we thank the other members of the Melbourne Children’s Working Group on Generative Artificial Intelligence in Child Health.

## Appendix 1. Survey questions

### Large Language Model Survey

**On behalf of the Melbourne Children’s Campus Working Group on Large Language Models**

We invite you to help us in some early discovery work on the current and potential future use of *Large Language Models* on our campus.

*Large Language Models* include tools such as Open AI ChatGPT (or other tools that use the ChatGPT engine); Google Bard; Microsoft Bing Chat; Jasper.ai and Perplexity.

Currently the campus partners have no policies/ positions on the use of these tools. We are aware that some tools are in use already – mostly by staff who are trying them out but also some more intensive use. We recognise that there may be both benefits and risks in using the methods.

You can answer this survey anonymously or provide your contact details if you wish for any follow ups.

***This survey will take 5-8 mins to complete***

#### Your role on Campus

* Which campus partner/s are you affiliated with?

□ RCH
□ MCRI
□ Uni Dept Paediatrics

* Which roles apply to your work on our campus?

□ Clinical work
□ Research
□ Teaching
□ Data Services for Campus Activities or Projects

Select all that apply

* Which best describes your level of experience?

□ Junior / trainee / student / junior data analyst or developer
□ Senior / Fully qualified / Post-doctoral / senior data analyst or developer

Please choose the response closest to your experience

#### Clinical

If you are not involved in clinical work, leave this page blank and click “Next”

**For Clinical Staff** - which craft group do you belong to?

□ Nursing
□ Allied Health
□ Medical
□ No answer

If you have used these tools for any clinical purpose, please provide some brief details of how and your experience so far:

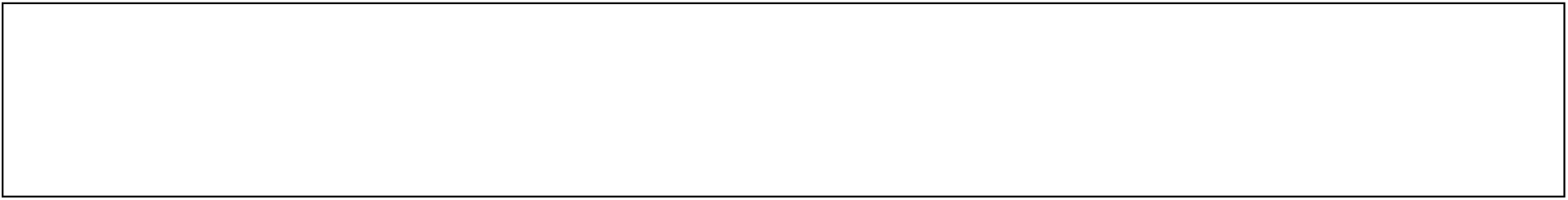

Examples might include: generating medical documentation, letters, patient/family education materials or other written output; suggesting diagnoses, investigations or treatments.

What future uses can you envisage for the use of these tools in clinical care?

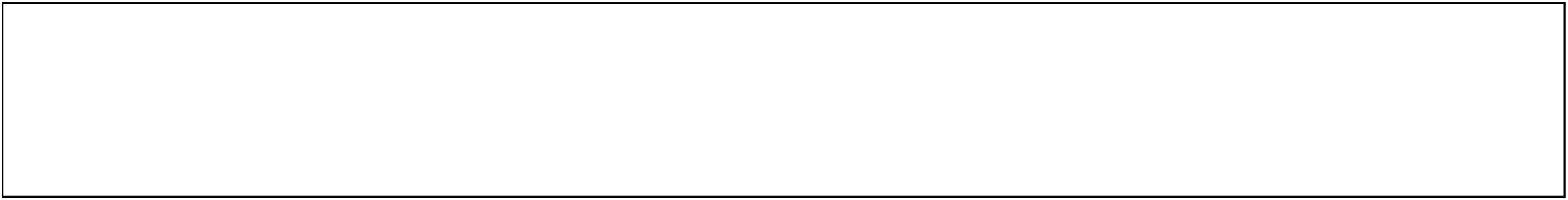

What do you consider to be the main opportunities and risks to using generative AI tools in clinical work?

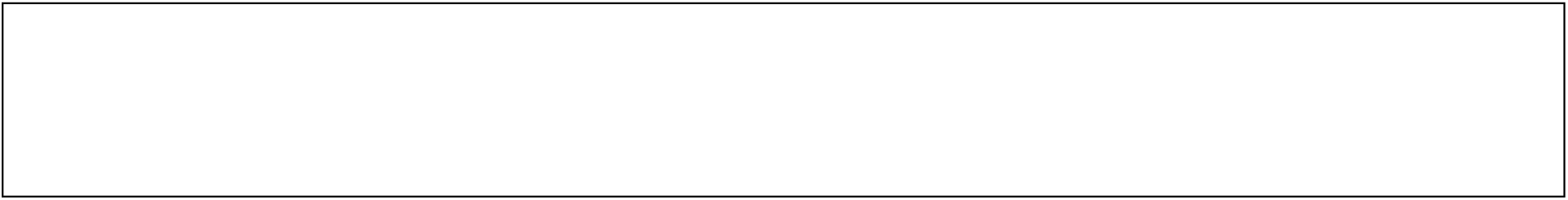

Examples include but not limited to: Opportunities: time saving, improved quality, productivity. Risks: accuracy, bias, privacy etc.

#### Teaching

If you are not involved in teaching, leave this page blank and click “Next”

If you have used these tools for any teaching purpose, please provide some brief details of how and your experience so far:

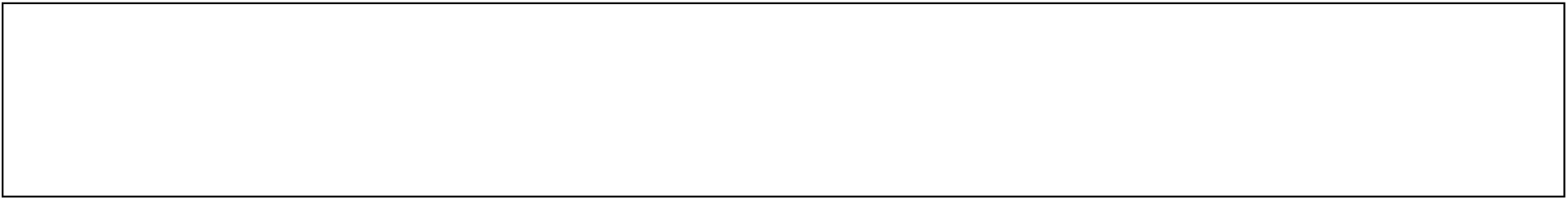

Examples might include as a student re-formatting or improving grammar in your assignments. As a teacher, encouraging students to use these tools for their submissions, generating multiple versions of assignment or quiz questions, generating images in lieu of stock images for slides.

What future uses can you envisage for the use of these tools in teaching and learning?

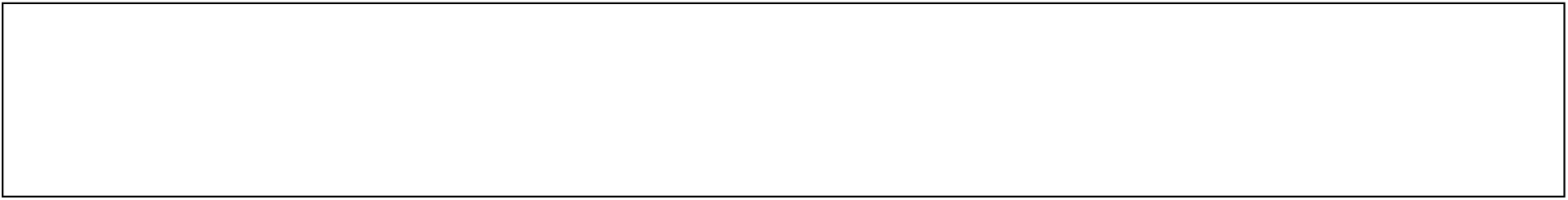

What do you consider to be the main opportunities and risks to using generative AI tools in research and research-related work?

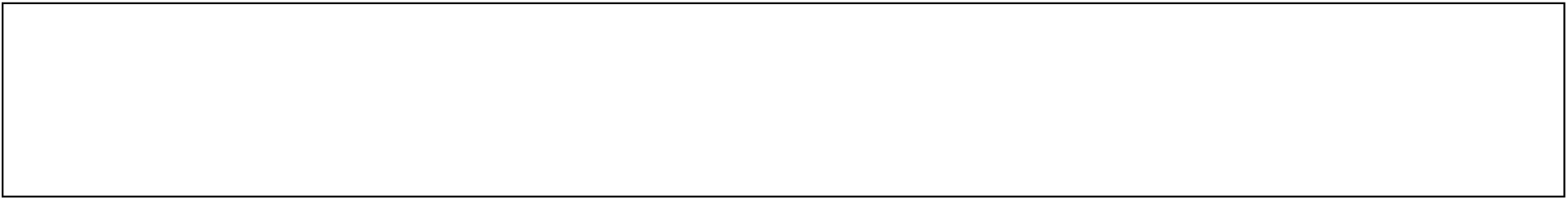

Examples include but not limited to: Opportunities: none, time saving, improved quality, productivity. Risks: none, accuracy, bias, privacy etc.

#### Research

If you are not involved in research, leave this page blank and click “Next”

For research staff - which area do you belong to?

□ Research
□ Research support
□ Operation
□ Other:
□ No answer

If you have used open generative AI tools for any research-related purpose, please provide some brief details of what you used them for and your experience with using them so far.

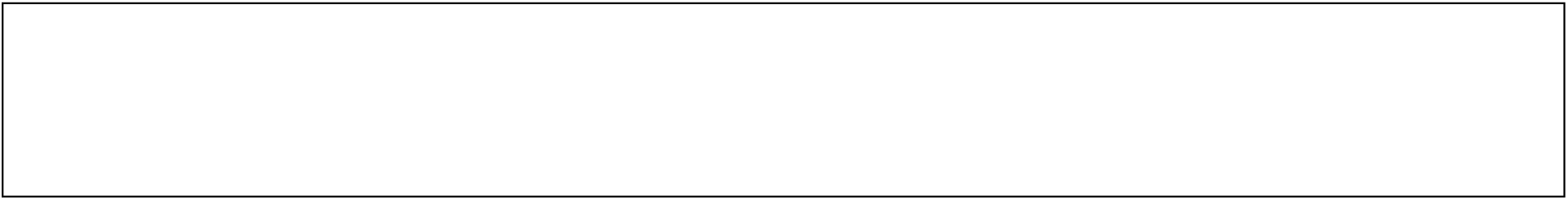

Examples might include re-formatting or improving grammar in your research, consent forms or other written outputs, developing code.

What future uses can you envisage for the use of these tools in your research and work-related environment?

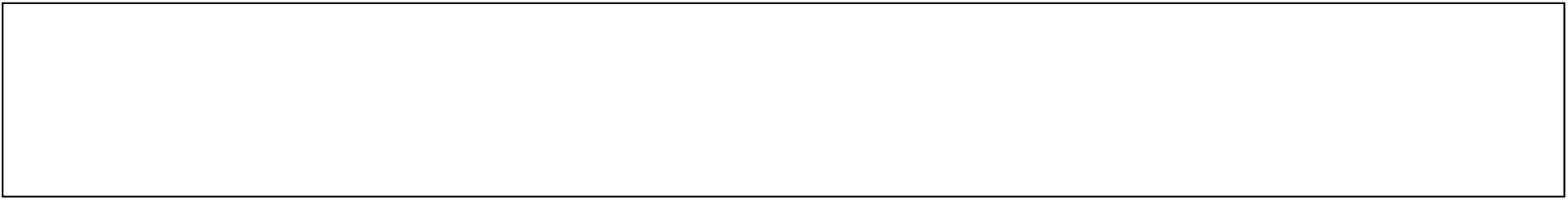

What do you consider to be the main opportunities and risks to using generative AI tools in research and research-related work?

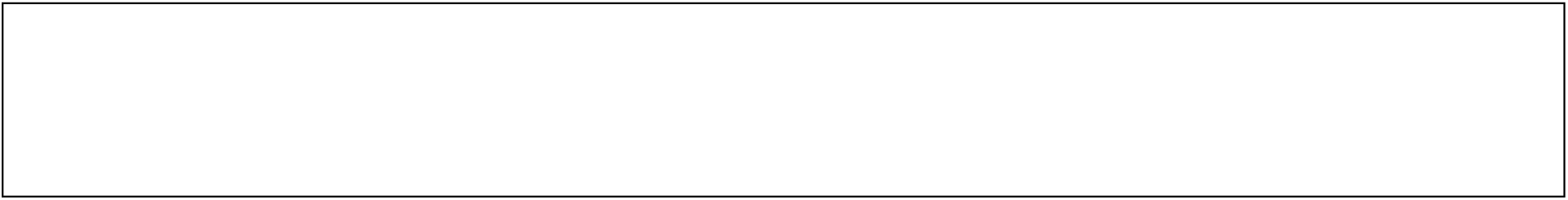

Examples include but not limited to: Opportunities: none, time saving, improved quality, productivity. Risks: none, accuracy, bias, privacy etc.

How do you use Large Language Models in your research?

□ As a core part of my research methods
□ To support my research
□ I’m not using these tools
□ Other:
□ No answer

Explanation

**As a core part of my research methods**

Examples include where large language models are a formal part of your data services methods (e.g., computational linguistics, natural language processing).

**To support my research**

Examples include re-formatting or improving grammar in my research, consent forms or other written outputs, developing code.

#### Tools

How familiar are you with these LLM tools?

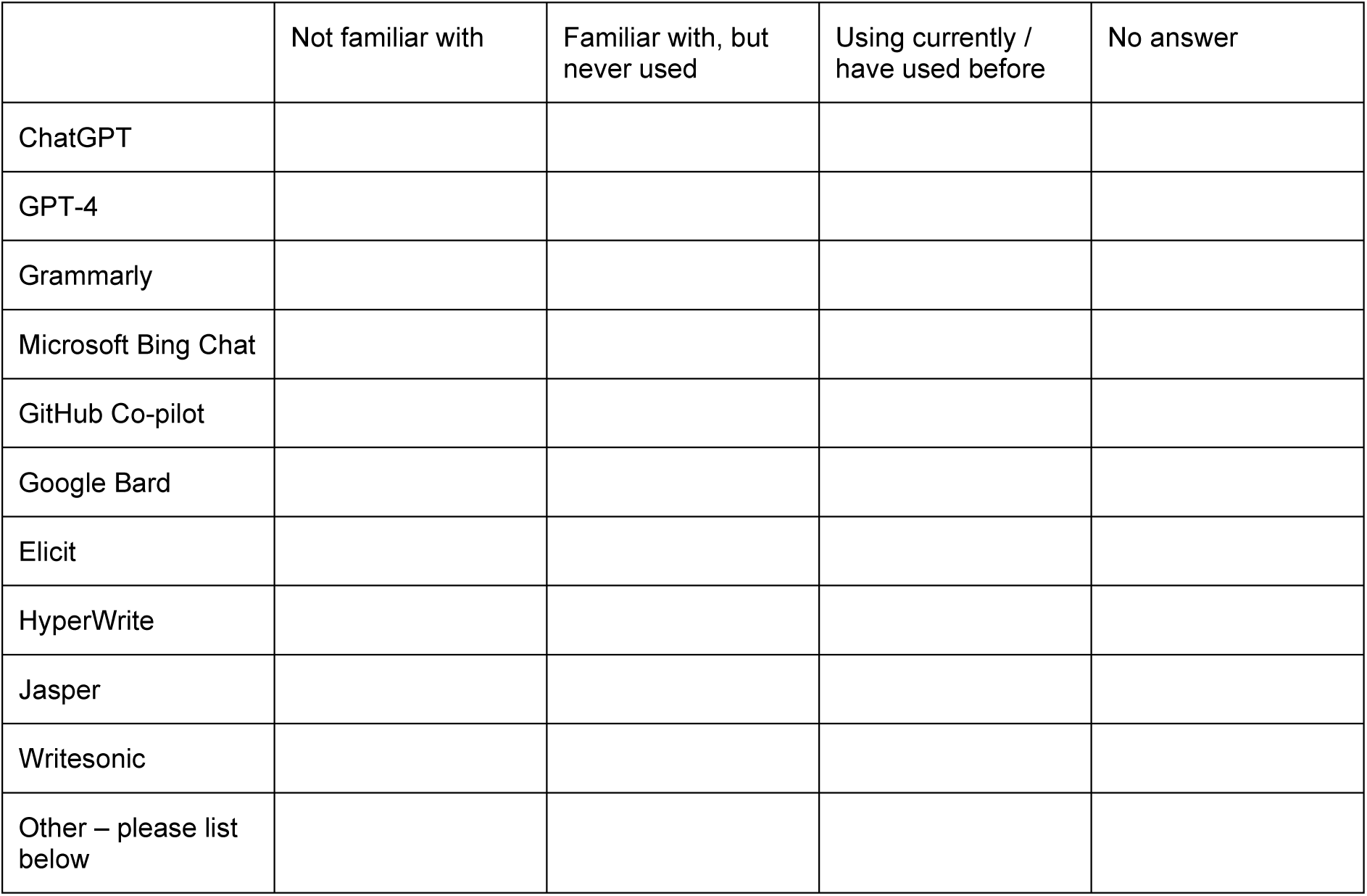

List other tools

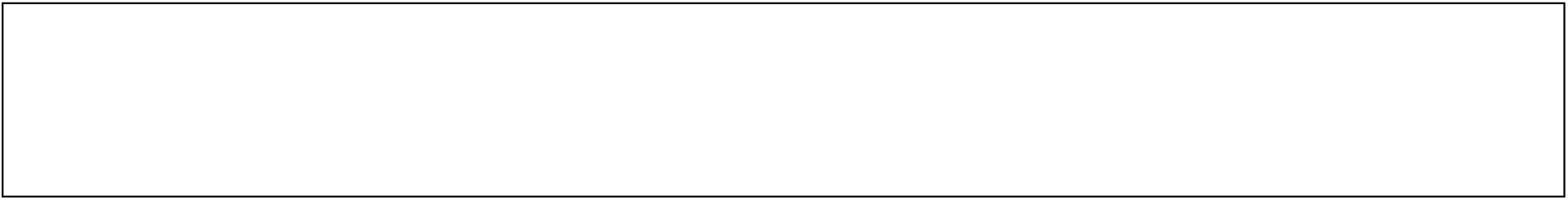

#### Your contact details

You can submit this survey anonymously (just click Submit) or leave your details if you would like to be involved in any follow ups.

Your name +/- position

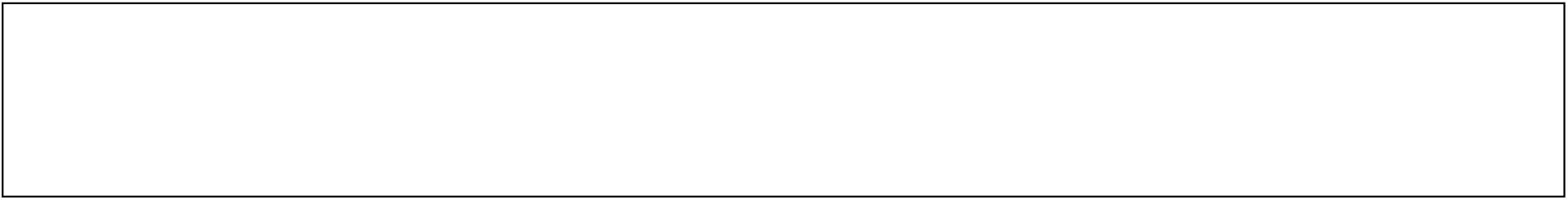

Your e-mail address

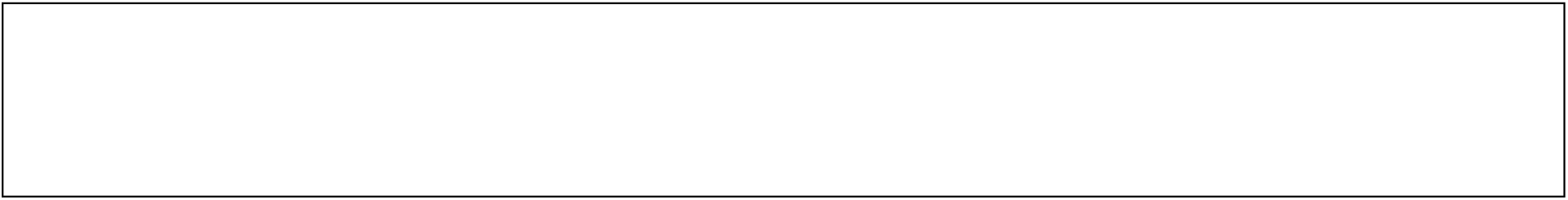

## Appendix 2. Recommendations for the use of Generative Artificial Intelligence (GenAI) and LLMs at our campus

**Table 1.**
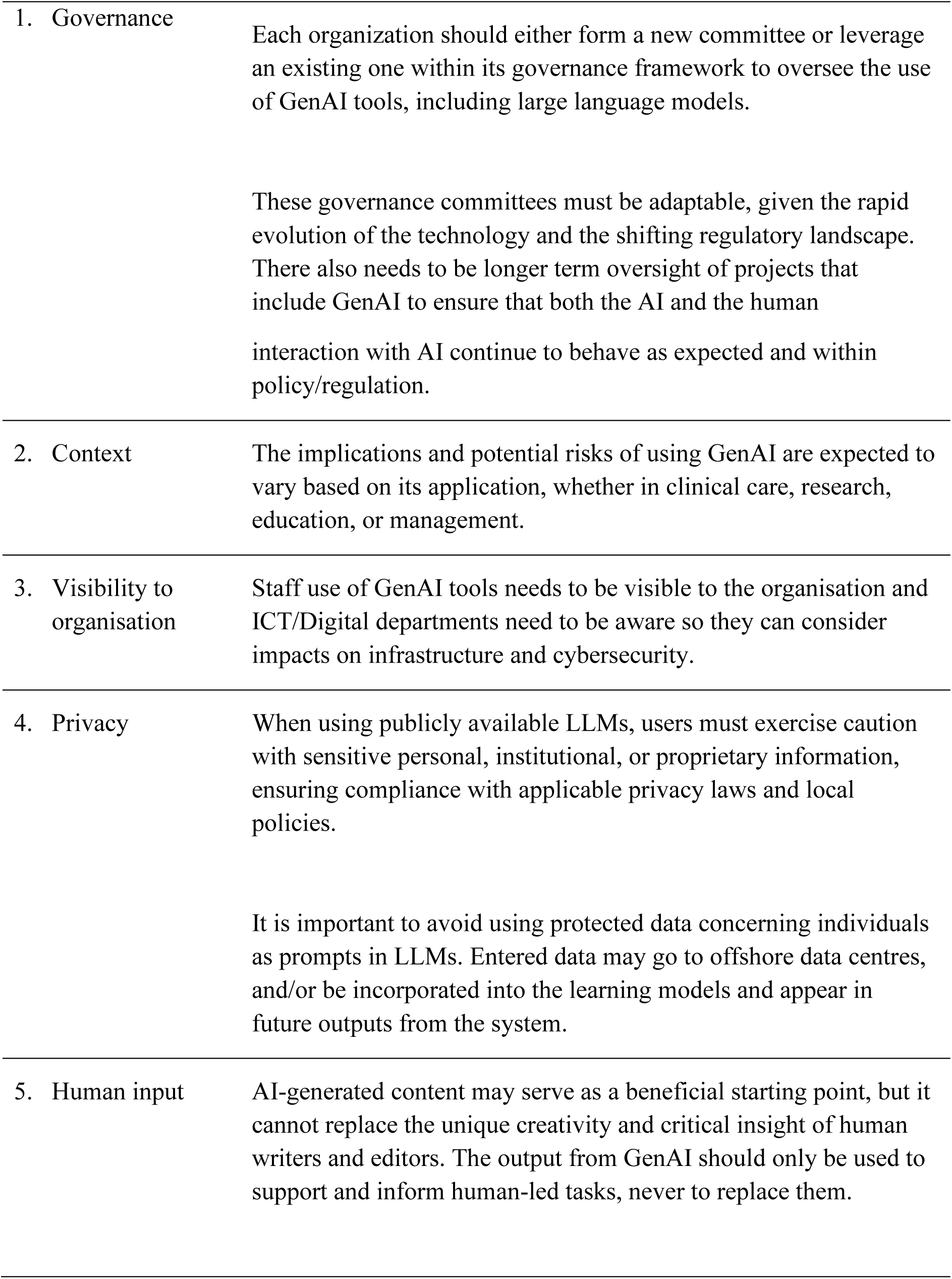
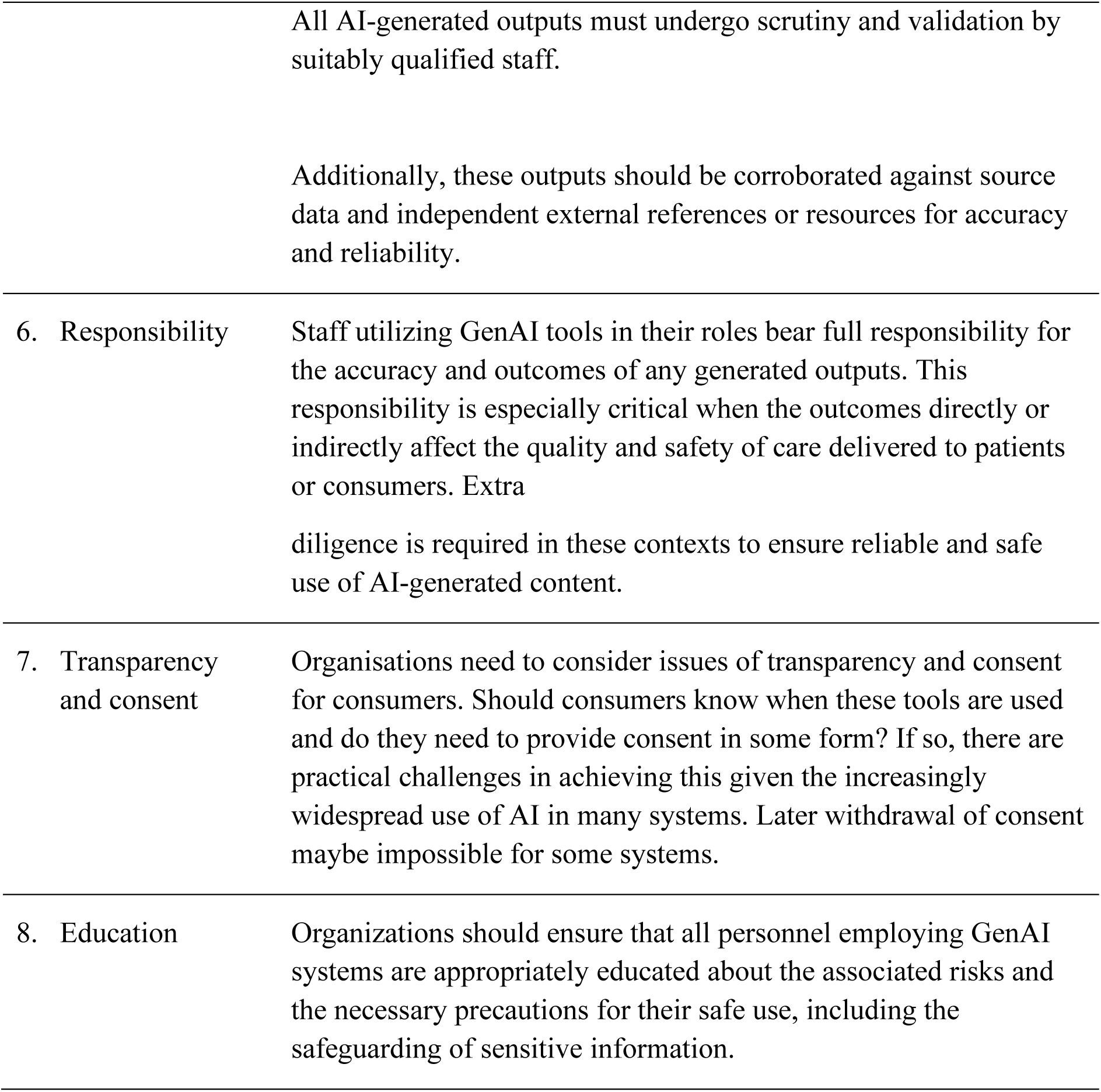
Recommendations for the use of Generative Artificial Intelligence (GenAI) and LLMs at our Campus.

## References

1. Naveed H, Khan AU, Qiu S, Saqib M, Anwar S, Usman M, et al. A Comprehensive Overview of Large Language Models [Internet]. arXiv; 2024 [cited 2024 May 14]. Available from: http://arxiv.org/abs/2307.06435

2. Bender EM, Koller A. Climbing towards NLU: On Meaning, Form, and Understanding in the Age of Data. In: Proceedings of the 58th Annual Meeting of the Association for Computational Linguistics [Internet]. Online: Association for Computational Linguistics; 2020 [cited 2023 Jul 12]. p. 5185–98. Available from: https://www.aclweb.org/anthology/2020.acl-main.463

3. Kasneci E, Sessler K, Küchemann S, Bannert M, Dementieva D, Fischer F, et al. ChatGPT for good? On opportunities and challenges of large language models for education. Learn Individ Differ. 2023 Apr;103:102274.

4. Sahoo SS, Plasek JM, Xu H, Uzuner Ö, Cohen T, Yetisgen M, et al. Large language models for biomedicine: foundations, opportunities, challenges, and best practices. J Am Med Inform Assoc. 2024 Apr 24;ocae074.

5. Barman KG, Wood N, Pawlowski P. Beyond transparency and explainability: on the need for adequate and contextualized user guidelines for LLM use. Ethics Inf Technol. 2024 Sep;26(3):47.

6. Zaretsky J, Kim JM, Baskharoun S, Zhao Y, Austrian J, Aphinyanaphongs Y, et al. Generative Artificial Intelligence to Transform Inpatient Discharge Summaries to Patient-Friendly Language and Format. JAMA Netw Open. 2024 Mar 11;7(3):e240357.

7. Abràmoff MD, Tarver ME, Loyo-Berrios N, Trujillo S, Char D, Obermeyer Z, et al. Considerations for addressing bias in artificial intelligence for health equity. Npj Digit Med. 2023 Sep 12;6(1):170.

8. Chin MH, Afsar-Manesh N, Bierman AS, Chang C, Colón-Rodríguez CJ, Dullabh P, et al. Guiding Principles to Address the Impact of Algorithm Bias on Racial and Ethnic Disparities in Health and Health Care. JAMA Netw Open. 2023 Dec 15;6(12):e2345050.

9. Safranek CW, Sidamon-Eristoff AE, Gilson A, Chartash D. The Role of Large Language Models in Medical Education: Applications and Implications. JMIR Med Educ. 2023 Aug 14;9:e50945.

10. Abd-alrazaq A, AlSaad R, Alhuwail D, Ahmed A, Healy PM, Latifi S, et al. Large Language Models in Medical Education: Opportunities, Challenges, and Future Directions. JMIR Med Educ. 2023 Jun 1;9:e48291.

11. Karabacak M, Margetis K. Embracing Large Language Models for Medical Applications: Opportunities and Challenges. Cureus [Internet]. 2023 May 21 [cited 2024 Aug 16]; Available from: https://www.cureus.com/articles/149797-embracing-large-language-models-for-medical-applications-opportunities-and-challenges

12. Lucas HC, Upperman JS, Robinson JR. A systematic review of large language models and their implications in medical education. Med Educ. 2024 Apr 19;medu.15402.

13. Rovenchak A, Druchok M. Machine learning-assisted search for novel coagulants: When machine learning can be efficient even if data availability is low. J Comput Chem. 2024 May 15;45(13):937–52.

14. Huang ETC, Yang JS, Liao KYK, Tseng WCW, Lee CK, Gill M, et al. Predicting blood–brain barrier permeability of molecules with a large language model and machine learning. Sci Rep. 2024 Jul 9;14(1):15844.

15. Butson R, Spronken-Smith R. AI and its implications for research in higher education: a critical dialogue. High Educ Res Dev. 2024 Apr 2;43(3):563–77.

16. O’Brien BC, Harris IB, Beckman TJ, Reed DA, Cook DA. Standards for Reporting Qualitative Research: A Synthesis of Recommendations. Acad Med. 2014 Sep;89(9):1245–51.

17. Writer. The state of generative AI in the enterprise [Internet]. Writer; 2024 [cited 2024 Aug 16]. Available from: https://writer.com/guides/generative-ai-survey/

18. Lumivero. NVivo [Internet]. 2023. Available from: www.lumivero.com

19. Victorian Department of Health. Health service use of unregulated Artificial Intelligence (AI) [Internet]. Victorian Department of Health; 2023 Jul. Available from: https://www.safercare.vic.gov.au/sites/default/files/2023-07/Advisory%20-%20ChatGPT%20and%20Generative%20AI%20July%202023%20FINAL.pdf

20. Epic Systems Corporation. Epic. 2024 [cited 2024 Sep 6]. Artificial Intelligence. Available from: https://www.epic.com/software/ai/

21. Department of Industry, Science and Resources. Safe and responsible AI in Australia [Internet]. Canberra, Australia: Australian Government; 2023 Jun. Available from: https://consult.industry.gov.au/supporting-responsible-ai

